# Prognostic impact of dynamic evolution in renal function among patients undergoing elective percutaneous coronary intervention

**DOI:** 10.1101/2023.02.03.23285431

**Authors:** Yifei Xiang, Xueqin Lin, Xiaoling Cai, Liwei Zhang, Manqing Luo, Jilang Zeng, Yansong Guo, Kai-Yang Lin

**Author notes:** **Correspondence:** Kai-yang Lin, Yan-song Guo. These authors have contributed equally to this work and share first authorship (Yifei Xiang, Xueqin Lin).

## Abstract

**Background:** Previous studies have shown that chronic kidney disease(CKD) affected the long-term prognosis of patients underwent the elective percutaneous coronary intervention(EPCI). However, the prognostic impact in patients with the development of the contrast-associated acute kidney injury(CA-AKI) and recovery or progression of CA-AKI were controversial. For the moment, little attention has been paid to the relationship between the dynamic evolution of renal function and its prognosis.

**Methods:** We used three stages to characterize the dynamic evolution of renal function, namely the occurrence of CKD at baseline, the occurrence of CA-AKI in the postoperative period and the occurrence of post kindey injury(PKI) at 3 - 6 months postoperatively. Cardiac death and all-cause mortality were used as the endpoint of the study. PKI(+) was defined as CA-AKI not recovered or an increase in absolute serum creatinine (SCr) ≥0.3 mg/dl or a SCr relative increase in creatinine ≥ 50% after 3 - 6 months. PKI(-) was defined as CA-AKI recovered or SCr elevation not meeting the PKI(+) requirement.

**Results:** We prospectively enrolled 2951 patients who underwent EPCI from 2012 to 2018. They were divided into three groups according to baseline CKD and CA-AKI: STAGE I[Unimpaired renal function group, CKD(-)/CA-AKI(-) (n=1247)], STAGE II[Partially impaired renal function group, IIa: CKD(-)/CA-AKI(+) (n=91) and IIb: CKD(+)/CA-AKI(-) (n=1472)] and STAGE III[severely impaired renal function group, CKD(+)/CA-AKI(+) (n=141)]. Subsequently, based on the occurrence of PKI, they were divided into six groups: STAGE I/PKI(-) (n=1212), STAGE I/PKI(+) (n=35), STAGE II/PKI(-) (n=1508), STAGE II/PKI(+) (n=55), STAGE III/PKI(-) (n=108), STAGE III/PKI(+) (n=33). In a mean follow-up period of 3.33± 1.39 years, we found that from STAGE I, STAGE II to STAGE III at baseline groups, the incidence of the primary outcome significantly increased. Meanwhile, from the baseline groups to the follow-up groups, the dynamic changes in renal function were observed. At the follow-uo groups, the occurrence of PKI did not affect the prognosis of patients in the STAGE I group(hazard ratio [HR] = 0.94, 95%CI: 0.15–8.11, p = 0.949) and the STAGE III group(hazard ratio [HR] = 1.19, 95%CI: 0.50–2.83 p = 0.689). However, for the STAGE II group (hazard ratio [HR] = 2.65, 95%CI: 1.42–4.94, p = 0.002), the development of PKI would lead to a poor prognosis for patients.

**Conclusion:** In patients undergoing EPCI, the occurrence of CKD and CA-AKI affected the long-term prognosis of patients. The prognostic impact of the occurrence of PKI depended on the renal function of patients. In patients with unimpaired renal function or severely impaired renal function, the prognostic impact of PKI was negligible. However, in patients with partially impaired renal function, avoidance of PKI could beneficial for their long-term prognosis.

## Introduction

Previous studies showed that patients undergoing PCI who have CKD have a higher mortality rate.^1^ This may be associated with their increased bleeding complications, peri-procedural myocardial infarction and in-stent thrombosis after PCI.^2^ This also demonstrates that CKD is associated with a worsening prognosis in patients with coronary artery disease (CAD).^3^

CA-AKI is one of the common complications after PCI and the third most common cause of acute kidney injury in hospitalized patients. In available studies, its incidence ranges from less than 5% in unselected populations to 50% in high-risk populations.^4-10^ The development of CA-AKI is strongly associated with increased length of stay, proportion of chronic renal failure, and poor prognosis.^11, 12^ Therefore, further exploration of the developmental process of CA-AKI is warranted.

CA-AKI is a typical example of reversible acute kidney injury.^13^ Recovery may occur early during acute kidney injury (AKI) up to 7 days after the insult, or later during acute kidney disease (AKD), between 7 days and 3 months after the insult to the kidney.^14^ However, when renal injury persists for more than 3 months, their renal function stabilizes, at which point the deterioration in renal function is irreversible.^14^ Therefore, we use PKI to define such state.

This is the dynamic evolution of renal function in patients after PCI, from their baseline renal function to post-operative CA-AKI to PKI after 3 months. However, current studies have not devoted much attention to the relationship between the dynamic evolution of renal function and prognosis. We aimed to examine the dynamic evolution of renal function and their prognostic impact in patients undergoing EPCI.

## Methods

The study followed the Helsinki Declaration principles and ethical approval was granted by the Fujian Provincial Hospital ethics committee (Ethical approval number: K2019–07–011). Written informed consent was obtained from all patients.

## Study population

This is a prospective observational study conducted at Fujian Provincial Hospital, Fujian Cardiovascular Institute, from January 2012 to December 2018. A total of 3030 patients who underwent EPCI were enrolled. The exclusion criteria were patients with long-term dialysis treatment, drug taking that decreasing kidney function 48h before CM exposure, including sodium bicarbonate, non-steroidal anti-inflammatory drugs (NSAIDs), metformin, aminoglycoside drugs, cyclosporine, cisplatin and so on and severe valvular heart disease or preparation for other operations. And finally 2951 patients were included in the study.

## Laboratory investigations, cardiac catheterization and medications

All patients’ clinical data were obtained from case records, including patients’ general conditions, laboratory tests, medication records, surgical data and cardiac ultrasound. SCr was measured at admission and daily for the 2 days after contrast exposure, as well as remeasured 3 to 6 months after discharge.

PCI was performed by experienced interventional cardiologists according to standard clinical practice using standard guide catheters, guide wires, balloon catheters and stents via the femoral or radial approach. All patients received nonionic, low-osmolar contrast media (either Iopamiron or Ultravist, both 370 mg I/ mL). In addition, 0.9% normal saline at a rate of 1 mL/kg/ h was administered intravenously approximately 12 hour during perioperative period (0.5 mL/kg/h if patients with heart failure).

The use of medications includs antiplatelet agents (aspirin/clopidogrel), β-adrenergic blocking agents, statins, angiotensin-converting enzyme inhibitor (ACEI) / angiotensin receptor blocker (ARB), and other drugs which were at the discretion of the cardiologists according to clinical protocols based on interventional guidelines.

### Definitions and follow-up

The primary endpoint was all-cause mortality, definded as death occurring after discharge from an EPCI procedure, excluding force majeure factors such as natural disasters and car accidents. The secondary endpoint was cardiac death, defined as any death due to cardiac cause.

CKD is defined as an estimated glomerular filtration rate(eGFR) <90 ml/min/1.73 m2 for 90 days.

The eGFR was calculated using the modified modification of diet in renal disease equation: 186.3 × SCr^-1.154^ × (age in years) ^-0.203^ × 1.212 (if patient was black) × 0.742 (if patient was female)

CA-AKI is defined as an increase in absolute serum creatinine (SCr) ≥0.3 mg/dl or a 48-hour serum relative increase in creatinine ≥ 50%.

PKI(+) was defined as CA-AKI not recovered or an increase in absolute serum creatinine (SCr) ⩾0.3 mg/dl or a SCr relative increase in creatinine ⩾ 50% after 3 - 6 months.

PKI(-) was defined as CA-AKI recovered or SCr elevation not meeting the PKI(+) requirement.

All patients were subject to follow-up for more than three months. Follow-up events were carefully monitored and recorded by trained nurses using either outpatient clinical visits or telephone contact with the patients or their relatives after discharge.

### Statistical analysis

All data were analyzed with R4.1.2. We compared the baseline characteristics among 4 groups divided by CKD and CA-AKI. Normally distributed continuous variables are expressed as mean + standard deviation (SD). The Student’s t-test, Wilcoxon rank sum test or one way-analysis of variance was performed to determine the differences among groups. Categorical variables were compared by chi-square test or Fisher exact test. Multivariate Cox regression analyses were used to identify the relationship between the dynamic evolution of CA-AKI and long-term prognosis. The Kaplan-Meier curve was used to assess the survival time between different groups. A 2-sided p value <0.05 was considered significant.

## Results

### The dynamic evolution of renal function and grouping

As shown in **FIGURE1A**, from CKD, CA-AKI, AKD to PKI, this was dynamic evolution of renal function in patients undergoing EPCI. However, over time, CA-AKI and AKD are subject to a variety of conditions such as early recovery, late recovery, and no recovery. Accordingly, as shown in **FIGURE1B**, we divided them into baseline, postoperative and 90 days postoperative.

### Baseline characteristics

This study included 2951 patients, they were divided into four groups based on renal function as reflected by the CKD and CA-AKI. As the stage rised, we could identify that the STAGE III group had more pathological conditions such as hypertension, diabetes and atrial fibrillation, poorer physical conditions such as lower left ventricular ejection fraction and worse cardiac function, worse laboratory datas such as higher BNP_baseline, higher SCR_baseline, higher URI_baseline and lower HGB_baseline. The prognosis of patients varies by groups.(**TABLE1**)

**TABLE1.**
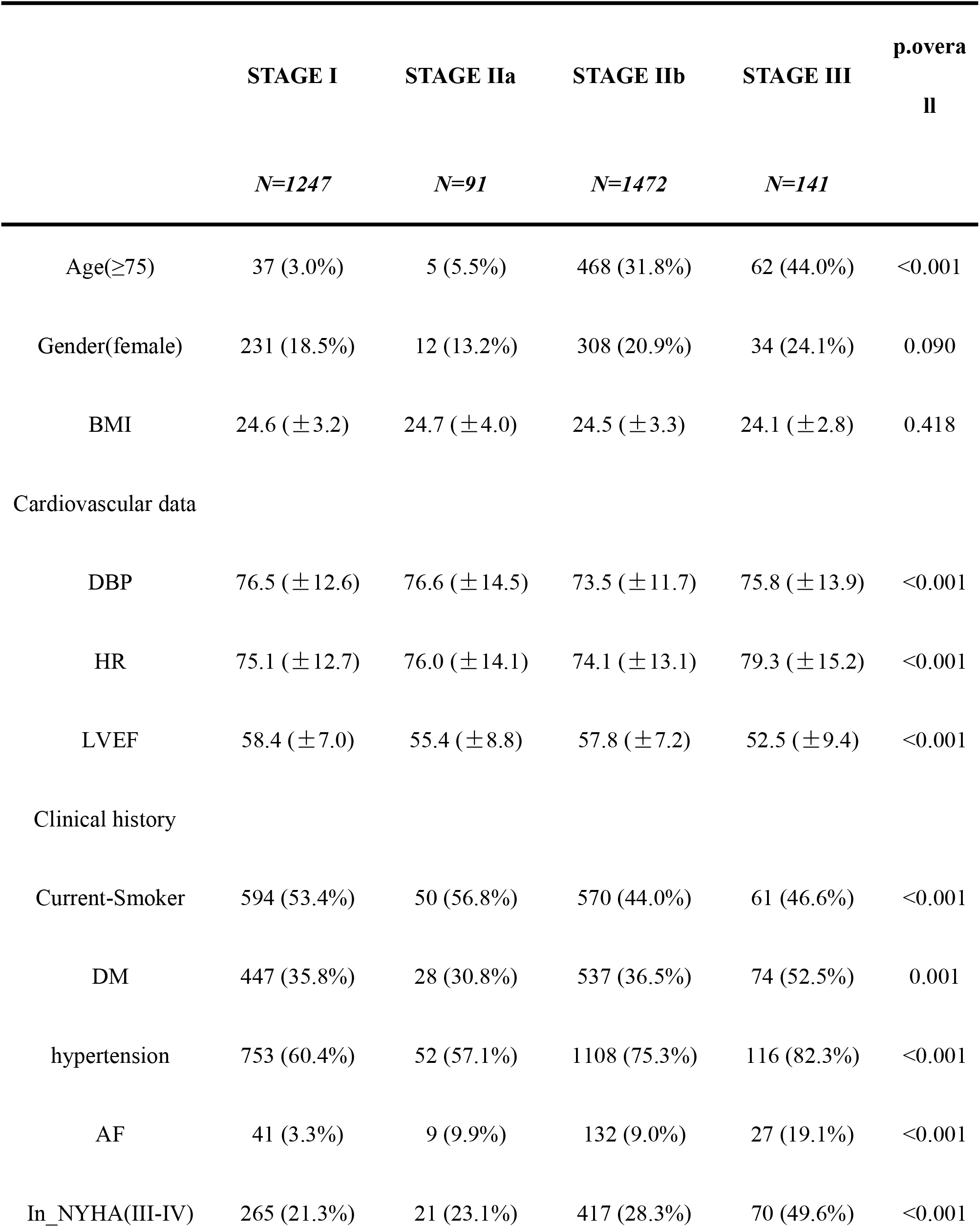
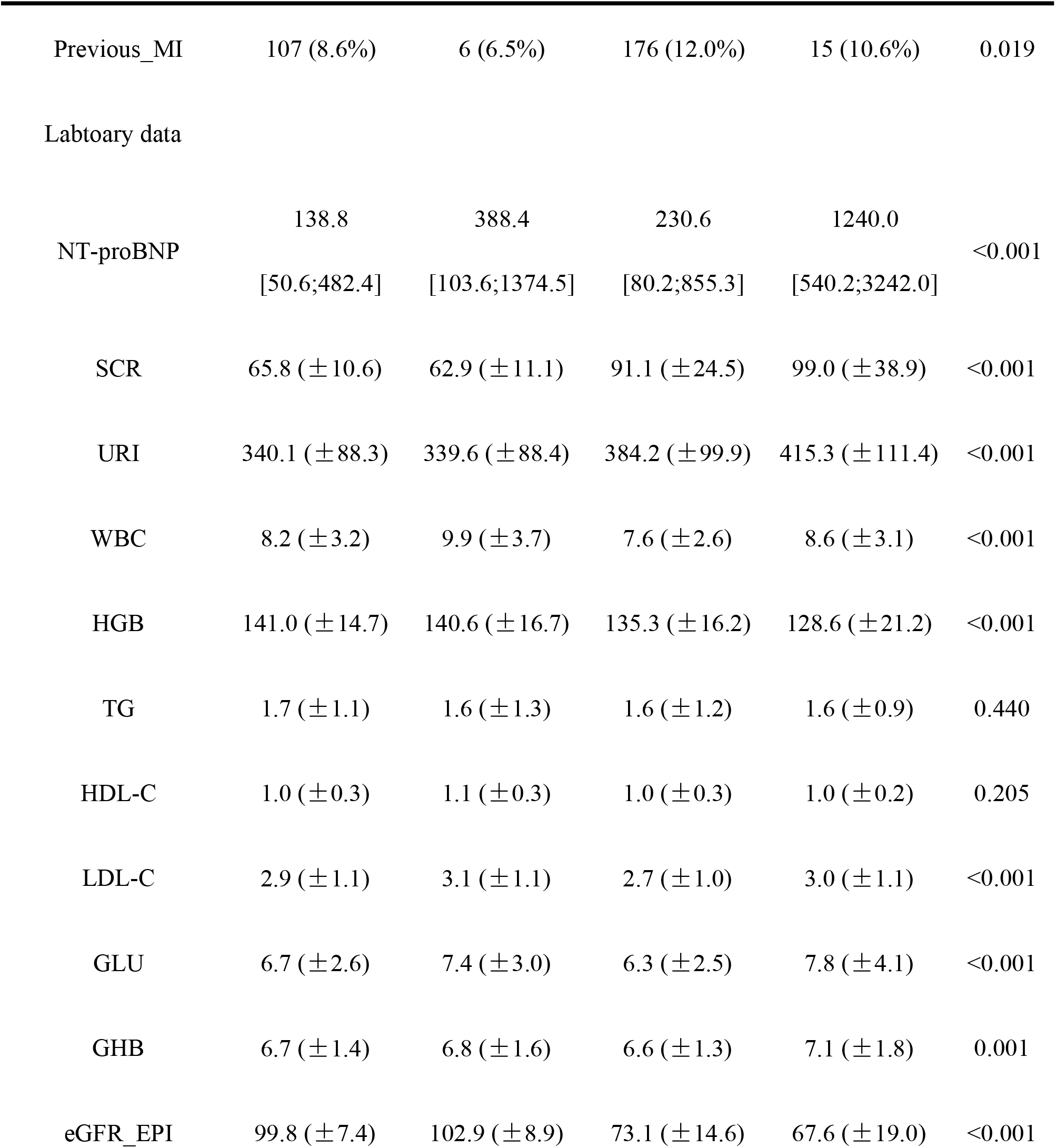
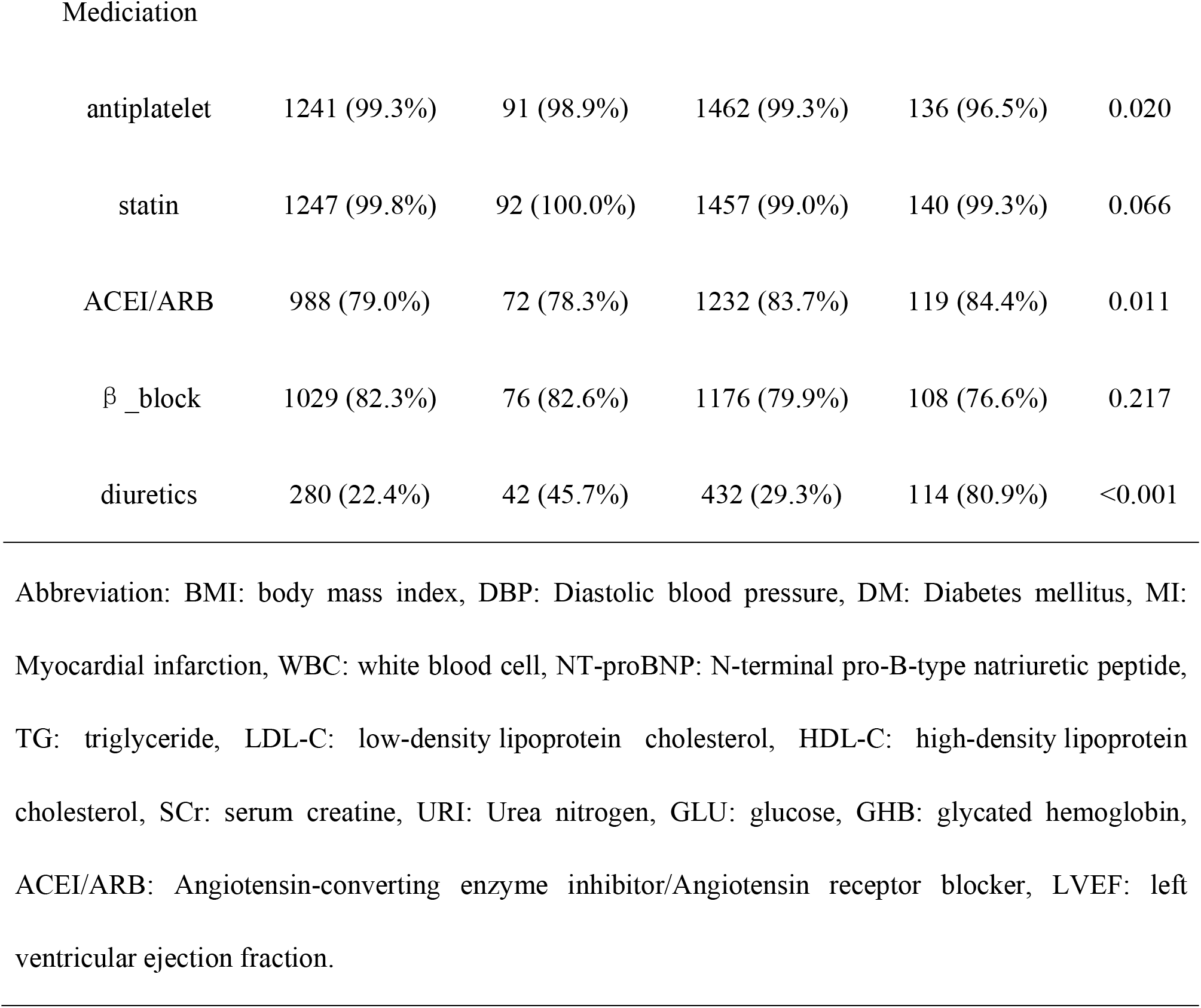
Baseline characteristics of the three groups according to CKD and CA-AKI.

### Baseline group and long-term prognosis

Accoring to the **FIGURE2**, the Kaplan-Meier curve showed that the different effects of each group on long-term prognosis, with the STAGE III group more likely to have a poorer long-term prognosis. However, we also identified that the survival curves of the two groups, STAGE IIIa and STAGE IIIb, were nearly overlapping, indicating that the prognostic effects of both were roughly equivalent, and therefore these two groups were classified into the STAGE II group (**Supplementary FIGURE1A**). This trend could indicate that an aggravation of renal impairment could affect the long-term prognosis of patients. And when the endpoint was set to cardiac death, the results obtained were consistent with the all-cause mortality (**FIGURE2B**).

In the **FIGURE3**, after adjusting for other risk factors in Model 2, Cox regression analysis for the STAGE II(hazard ratio [HR] = 2.19 95%CI: 1.44, 3.30, p< 0.001) and STAGE III groups (hazard ratio [HR] = 3.74, 95%CI: 2.18, 6.42, p< 0.001) indicated that both CKD and CA-AKI affected the long-term prognosis of patients. Similarly, in the Cox regression analysis with cardiac death as the endpoint, CKD and CA-AKI still influenced the long-term prognosis of patients.

### Follow-up group and long-term prognosis

According to the **Supplementary FIGURE2A**, the impact of the occurrence of PKI on the 5-year survival of patients was influenced by renal function, with STAGE III/PKI(+) having the highest mortality rate, followed by STAGE II/PKI(+). This suggested that for the STAGE II group, the occurrence of PKI severely affected the survival rate of patients. Based on the **Supplementary FIGURE2B**, similar results to those described above were obtained by setting cardiac death as the endpoint.

In the follow-up group, after adjusting for other risk factors in model 2, we identified that for the STAGE I group (hazard ratio [HR] = 1.13, 95%CI: 0.15, 8.25, p = 0.907), the occurrence of PKI did not affect the long-term prognosis of patients. However, for the other groups, the prognostic impact of PKI was not well defined. (**FIGURE4**)

### Dynamic renal function changes in patients undergoing EPCI

Among 2951 patients who underwent EPCI, their Scr were measured at both 2days and 3 to 6 months. According to the **FIGURE5A**, we found that dynamic changes in renal function were observed in patients from the baseline groups to the follow-up groups. The transition rate from the transition rates from STAGE I to STAGE I/PKI(+) (2.806%), from STAGE IIa to STAGE IIa/PKI(+) (14.286%), from STAGE IIb to STAGE IIb/PKI(+) (2.853%), from STAGE III to STAGE III/PKI(+) (23.404%), respectively. Based on the **FIGURE5A**, we found that the group that developed CA-AKI in the postoperative period had a higher chance of developing PKI. Among them, the STAGE III group had the highest rate of development into a PKI.

As time progresses, renal function can deteriorate or improve in patients after EPCI. Therefore, we compared the differences between the 5-year cumulative mortality between the different subgroups. Based on the **FIGURE5B**, we found that the 5-year cumulative mortality would increased as the renal function deteriorated. At the same time, we observed that for the STAGE II group, the occurence of the PKI significantly increased the 5-year cumulative mortality of patients. However, in the other groups, the phenomenon was not remarkable.

The results mentioned above were further validated in the **FIGURE5C**. In our study, we identified that for the STAGE I group (hazard ratio [HR] = 0.94, 95%CI:0.15–8.11, p = 0.949), and the STAGE III group (hazard ratio [HR] = 1.19, 95%CI:0.50–2.83 p = 0.689), the dynamic evolution of its CA-AKI outcome did not affect the long-term prognosis of patients. However, for the STAGE II group (hazard ratio [HR] = 2.65, 95%CI:1.42–4.94, p = 0.002), the STAGE II/PKI(+) group had a worse prognosis than the STAGE II/ PKI(-) group.

## Discussion

To our knowledge, this was the first study to investigate the relationship between the dynamic evolutionary process of renal function and prognosis in a population undergoing the EPCI.

In the article, The definition of PKI adopted for the study was developed by referring to the literature of Forni LG et al.^14^ This is a definition that focuses more on persistent impairment of renal function and does not take into account the patient’s underlying renal function. Because the existing definition of CKD, although also reflecting sustained impairment of renal function. This definition was mainly applied to population with normal renal function at baseline. In population with poor underlying renal function or progressive deterioration of renal function, there is no appropriate definition to clarify their persistent renal impairment. Therefore, we used PKI to define renal impairment beyond 3 months.

The main finding of this study was that CA-AKI and CKD affect the long-term prognosis of patients. And for the STAGE I and the STAGE III groups, there was no impact on the long-term prognosis of patients regardless of the occurrence of PKI. In contrast, for the STAGE II group, the occurrence of PKI resulted in a poor long-term prognosis for the patients. This suggested that the impact of PKI on the long-term prognosis of patients was based on the renal function of patients. In patients with unimpaired renal function or severely impaired renal function, the development of PKI did not affect their long-term prognosis.

According to the **FIGURE6A**, we can identify a linear relationship between eGFR and all-cause mortality, with a gradual increase in all-cause mortality in patients as eGFR decreased. As reported in previous articles, the development of CKD was an independent risk factor for patient death and also leads to a poor prognosis for patients. And it can have more serious consequences as the stage of CKD increased.^15, 16^ This may be related to worse underlying condition of patients with CKD. As shown in the **FIGURE6B**, we found a U-shaped relationship between eGFR and CA-AKI, suggesting a controversial relationship between CA-AKI and prognosis. However, according to our findings, CA-AKI does affect the long-term prognosis of patients. The occurrence of CA-AKI had been shown in previous studies to affect long-term prognosis of patients. This coincides with our results. The results may be related to the inflammatory state, hyperglycemic state, elevated D-dimers, and elevated NT-ProBNP that occur after the onset of CA-AKI.^17-20^

Previous studies had shown that patients with nomal renal function who underwent PCI did not have a significant impact on patient prognosis.^21^ In addition, according to the available studies, contrast medium (CM) caused less damage in patients with normal renal function.^22^ Therefore, for patients with normal renal function, even if CA-AKI occurs, its effect on renal function was negligible. And, according to **Supplementary FIGURE2**, we found that the occurrence of PKI did not make a large difference in patient survival for the STAGE I group. In summary, the occurrence of PKI did not affect the long-term prognosis of patients with normal renal function.

According to the **FIGURE2** and **Supplementary FIGURE2**, we can identify that the STAGE III group has the worst prognosis in the baseline groups. Likewise, the prognosis of the STAGE III/PKI(+) group was also particularly poor in the follow-up groups. Howwver, based on **FIGURE5C**, even if patients in the STAGE III group showed recovery of renal function, it was not meaningful for their long-term prognosis. The presence of this outcome requires taking end-stage renal disease(ESRD) into account. According to the literature of Forni LG et al, for CKD patients with cardiovascular disease, the occurrence of CA-AKI can lead to the ESRD.^14^ This is considered to be caused by inflammation, fibrosis and vascular rarefaction leading to persistent cell and tissue malfunction that occurs in patients with CKD after the onset of CA-AKI.

According to **FIGURE5B**, we can identify that for the STAGE II group, the ocurence of PKI would lead to an increase in the incidence of endpoint events. And in accordance with **FIGURE5C**, the prognosis of the STAGE II/PKI(+) group was worse than that of the STAGE II/PKI(-) group. This was consistent with the results reported by JIN LIU et al. This result may be caused by the occurrence of PKI that aggravates the patient’s existing CKD.^23^ As to why patients without AKI developed PKI, the reasons may be related to reduced renal perfusion or parenchymal kidney disease in which the decline in eGFR is too small or progresses too slowly to elevate Scr within the time limits of AKI criteria, or by infrequent Scr measurements to detect.^24^ This could be used to explain why some patients can progress to PKI in the absence of CA-AKI.

Our results suggested that for patients undergoing EPCI, it was important to first assess their renal function. When a patient had CKD, it was important to avoid the occurence of CA-AKI, otherwise the long-term prognosis of the patient was seriously affected. Even if CA-AKI did not occur, it was important to focus on the dynamic evolution of the patient’s renal function to avoid the development of PKI. In patients with normal renal function, the onset of CA-AKI was also to be avoided, as it can affect the long-term prognosis. However, the outcome of the dynamic evolution of renal function in such patients had little impact on the prognosis.

## Limitations

Our research has several limitations. Firstly, our study is a single-center prospective analysis, which may lead to a lack of representativeness of our results. However, our study has a large sample size, and the result has been validated in a multivariate logistic regression analyses, which makes our results more credible. Second, long-term mortality is complex and multivariable. However, we included both all-cause mortality and cardiac death as endpoints, and the results are worth promoting. Finally, the definition of CA-AKI is diverse. In our research, CA-AKI was defined as an increase in SCr ≤ 50% or ≤ 0.3 mg/d from baseline within 48h after the EPCI.

## Conclusion

In patients undergoing EPCI, the occurrence of CKD and CA-AKI affected the long-term prognosis of patients. The prognostic impact of the occurrence of PKI depended on the renal function of patients. In patients with unimpaired renal function or severely impaired renal function, the prognostic impact of PKI was negligible. However, in patients with partially impaired renal function, avoidance of PKI could beneficial for their long-term prognosis.

## Data Availability

Data available on request from the authors The data that support the findings of this study are available from the corresponding author, [Kai-Yang Lin], upon reasonable request.

## References

1. Liao GZ, Li YM, Bai L, Ye YY, Peng Y. Revascularization vs. Conservative Medical Treatment in Patients With Chronic Kidney Disease and Coronary Artery Disease: A Meta-Analysis. Front Cardiovasc Med. 2021;8:818958.

2. Sattar S, Ahmed N, Akhter Z, Aijaz S, Lakhani S, Malik R, Pathan A. In-Hospital outcomes in acute coronary syndrome patients with concomitant severe chronic kidney disease undergoing percutaneous coronary intervention. Pak J Med Sci. 2019;35:291–297.

3. Lima EG, Charytan DM, Hueb W, de Azevedo D, Garzillo CL, Favarato D, Linhares Filho J, Martins EB, Batista DV, Rezende PC, Hueb AC, Ramires J, Kalil Filho R. Long-term outcomes of patients with stable coronary disease and chronic kidney dysfunction: 10-year follow-up of the Medicine, Angioplasty, or Surgery Study II Trial. Nephrol Dial Transplant. 2020;35:1369–1376.

4. Mehran R, Aymong ED, Nikolsky E, Lasic Z, Iakovou I, Fahy M, Mintz GS, Lansky AJ, Moses JW, Stone GW, Leon MB, Dangas G. A simple risk score for prediction of contrast-induced nephropathy after percutaneous coronary intervention: development and initial validation. J Am Coll Cardiol. 2004;44:1393–1399.

5. Dangas G, Iakovou I, Nikolsky E, Aymong ED, Mintz GS, Kipshidze NN, Lansky AJ, Moussa I, Stone GW, Moses JW, Leon MB, Mehran R. Contrast-induced nephropathy after percutaneous coronary interventions in relation to chronic kidney disease and hemodynamic variables. Am J Cardiol. 2005;95:13–19.

6. Aspelin P, Aubry P, Fransson SG, Strasser R, Willenbrock R, Berg KJ. Nephrotoxic effects in high-risk patients undergoing angiography. N Engl J Med. 2003;348:491–499.

7. Briguori C, Colombo A, Airoldi F, Violante A, Castelli A, Balestrieri P, Paolo Elia P, Golia B, Lepore S, Riviezzo G, Scarpato P, Librera M, Focaccio A, Ricciardelli B. N-Acetylcysteine versus fenoldopam mesylate to prevent contrast agent-associated nephrotoxicity. J Am Coll Cardiol. 2004;44:762–765.

8. Mehta RL, Kellum JA, Shah SV, Molitoris BA, Ronco C, Warnock DG, Levin A. Acute Kidney Injury Network: report of an initiative to improve outcomes in acute kidney injury. Crit Care. 2007;11:R31.

9. Kolonko A, Kokot F, Wiecek A. Contrast-associated nephropathy--old clinical problem and new therapeutic perspectives. Nephrol Dial Transplant. 1998;13:803–806.

10. Nikolsky E, Mehran R, Turcot D, Aymong ED, Mintz GS, Lasic Z, Lansky AJ, Tsounias E, Moses JW, Stone GW, Leon MB, Dangas GD. Impact of chronic kidney disease on prognosis of patients with diabetes mellitus treated with percutaneous coronary intervention. Am J Cardiol. 2004;94:300–305.

11. Rihal CS, Textor SC, Grill DE, Berger PB, Ting HH, Best PJ, Singh M, Bell MR, Barsness GW, Mathew V, Garratt KN, Holmes DR Jr. Incidence and prognostic importance of acute renal failure after percutaneous coronary intervention. Circulation. 2002;105:2259–2264.

12. Gruberg L, Mintz GS, Mehran R, Gangas G, Lansky AJ, Kent KM, Pichard AD, Satler LF, Leon MB. The prognostic implications of further renal function deterioration within 48 h of interventional coronary procedures in patients with pre-existent chronic renal insufficiency. J Am Coll Cardiol. 2000;36:1542–1548.

13. Chawla LS, Bellomo R, Bihorac A, Goldstein SL, Siew ED, Bagshaw SM, Bittleman D, Cruz D, Endre Z, Fitzgerald RL, Forni L, Kane-Gill SL, Hoste E, Koyner J, Liu KD, Macedo E, Mehta R, Murray P, Nadim M, Ostermann M, Palevsky PM, Pannu N, Rosner M, Wald R, Zarbock A, Ronco C, Kellum JA. Acute kidney disease and renal recovery: consensus report of the Acute Disease Quality Initiative (ADQI) 16 Workgroup. Nat Rev Nephrol. 2017;13:241–257.

14. Forni LG, Darmon M, Ostermann M, Oudemans-van Straaten HM, Pettilä V, Prowle JR, Schetz M, Joannidis M. Renal recovery after acute kidney injury. Intensive Care Med. 2017;43:855–866.

15. Watabe H, Sato A, Hoshi T, Takeyasu N, Abe D, Akiyama D, Kakefuda Y, Nishina H, Noguchi Y, Aonuma K. Association of contrast-induced acute kidney injury with long-term cardiovascular events in acute coronary syndrome patients with chronic kidney disease undergoing emergent percutaneous coronary intervention. Int J Cardiol. 2014;174:57–63.

16. Silvain J, Nguyen LS, Spagnoli V, Kerneis M, Guedeney P, Vignolles N, Cosker K, Barthelemy O, Le Feuvre C, Helft G, Collet JP, Montalescot G. Contrast-induced acute kidney injury and mortality in ST elevation myocardial infarction treated with primary percutaneous coronary intervention. Heart. 2018;104:767–772.

17. You ZB, Lin KY, Zheng WP, Lin CJ, Lin F, Guo TL, Zhu PL, Guo YS. Association of prealbumin levels with contrast-induced acute kidney injury in elderly patients with elective percutaneous coronary intervention. Clin Interv Aging. 2018;13:641–649.

18. Lin KY, Wu ZY, You ZB, Zheng WP, Lin CJ, Jiang H, Ruan JM, Guo YS, Zhu PL. Pre-Procedural N-Terminal Pro-B Type Natriuretic Peptide Predicts Contrast-Induced Acute Kidney Injury and Long-Term Outcome in Elderly Patients After Elective Percutaneous Coronary Intervention. Int Heart J. 2018;59:926–934.

19. Lin KY, Shang XL, Guo YS, Zhu PL, Wu ZY, Jiang H, Ruan JM, Zheng WP, You ZB, Lin CJ. Association of Preprocedural Hyperglycemia With Contrast-Induced Acute Kidney Injury and Poor Outcomes After Emergency Percutaneous Coronary Intervention. Angiology. 2018;69:770–778.

20. Lin KY, Chen HC, Jiang H, Wang SY, Chen HM, Wu ZY, Jiang F, Guo YS, Zhu PL. Predictive value of admission D-dimer for contrast-induced acute kidney injury and poor outcomes after primary percutaneous coronary intervention. BMC Nephrol. 2020;21:90.

21. Kim TO, Kang DY, Ahn JM, Kim SO, Lee PH, Lee J, Kim JH, Kim HJ, Kim JB, Choo SJ, Chung CH, Lee JW, Park SJ, Park DW. Prognostic Impact of Mildly Impaired Renal Function in Patients Undergoing Multivessel Coronary Revascularization. J Am Coll Cardiol. 2022;79:1270–1284.

22. Li Q, Pan S. Contrast-Associated Acute Kidney Injury: Advances and Challenges. Int J Gen Med. 2022;15:1537–1546.

23. Liu J, Li Q, Chen W, Huang H, Yu Y, Wang B, Liang G, Lai W, Liu L, Ying M, Wei H, Huang Z, Ni J, Chen J, Chen S, Liu Y. Incidence and mortality of acute kidney disease following coronary angiography: a cohort study of 9223 patients. Int Urol Nephrol. 2022.

24. Chu R, Li C, Wang S, Zou W, Liu G, Yang L. Assessment of KDIGO definitions in patients with histopathologic evidence of acute renal disease. Clin J Am Soc Nephrol. 2014;9:1175–1182.

